# Evaluation of the performance and feasibility of RLDT in detecting *Shigella* in a primary healthcare facility of rural Bangladesh

**DOI:** 10.64898/2025.12.08.25341873

**Authors:** Sampa Dash, Eva Sultana, Kiely Flynn, Farina Naz, Mohammad Ali, Md. Razibur Rahman, Md. Mizanur Rohman, Shaniram Das, Tahmeed Ahmed, ASG Faruque, Subhra Chakraborty

## Abstract

Shigellosis remains underdiagnosed due to the lack of rapid, reliable diagnostics. Although *Shigella* spp. is known for causing dysentery, over 50% of *Shigella*-associated cases are watery diarrhea. Identifying and treating these cases of watery diarrhea caused by *Shigella* could be lifesaving, which would require a point-of-care (POC) *Shigella* test. Evidence-based treatment could also reduce the overuse of antibiotics. We evaluated the feasibility and applicability of the Rapid LAMP-based Diagnostic Test (RLDT) assay for detecting shigellosis in a healthcare facility. Stool samples (n=228) were collected from children seeking facility care during diarrhea and on their follow-up visits from an ongoing case-control study, INSIGHT. The stool samples were tested by the INSIGHT lab personnel using the RLDT for *Shigella* spp. The lab personnel at a rural primary healthcare facility, Mirzapur Upazila Health Complex (MUHC) in Bangladesh, received training in RLDT and retested the stool samples with RLDT at the MUHC; the results were compared. The acceptance of RLDT at MUHC was also evaluated through questionnaires. After training, the MUHC lab personnel independently performed RLDT. The RLDT tests performed by the MUHC showed sensitivity of 98% and specificity of 99%, with an almost perfect agreement (Kappa = 0.96) compared with the pre-tested RLDT results. The RLDT assay was well received by the MUHC, despite the general challenges of limited manpower and resources at rural health care facilities. This study demonstrates the potential of using RLDT as a POC test in *Shigella*-endemic countries to support evidence-based treatment, saving lives and reducing inappropriate antibiotic use.

**Keypoints:** RLDT, as a simple and accurate diagnostic tool, offers a practical solution for *Shigella* detection in low-resource settings. Scaling up RLDT will empower health systems by enabling timely diagnoses to guide treatment and reduce antimicrobial resistance.

## Introduction

*Shigella* species (*Shigella* spp.) *is* a primary cause of moderate-to-severe diarrhea (MSD) in children living in impoverished areas of the world. Although mortality has decreased in recent years to ∼70,000 [1, 2] there are at least 80 million cases of shigellosis each year with estimated disability adjusted life years (DALYS) at 7 million and year loss due to disability (YLDs) at 744,000 [1–4]. *Shigella* can present as dysentery or watery diarrhea, and may accompany complications like encephalopathy and rectal prolapse. *Shigella* infection could result in long-term adverse consequences, affecting linear growth, leading to stunting, and may delay psychomotor development [5–8]. WHO designates *Shigella* as a high-priority enteric pathogen for vaccine development [5, 6], although no licensed vaccine is yet available. Antimicrobial resistance of *Shigella* is rapidly increasing and currently poses a global threat. Shigellosis is one of the underdiagnosed and underreported diseases in low- and middle-income countries (LMICs), especially because of the absence of an appropriate diagnostic tool. The direct culture method is considered the gold standard diagnostic method for detecting *Shigella*. However, the culture method is less sensitive, requires 2-3 days, and is not often available in the health facilities or laboratories of LMICs [9]. Recent studies showed that analysis of stool samples using quantitative PCR revealed a significantly higher *Shigella*-associated diarrhea among children under 5 than previously estimated *Shigella*-attributed diarrhea cases using the culture method [9–12]. Conventional PCR and Quantitative PCR (qPCR) are highly sensitive but require a well-equipped laboratory facility and trained laboratory personnel, which are often only available at the referral laboratories and a few tertiary care hospitals of LMICs. Besides, these assays are expensive and require cold chain maintenance and, therefore, are unsuitable as point-of-care tests in low-resource settings where shigellosis is endemic.

Clinical symptoms, especially dysentery with visible blood in stool, are suggestive of shigellosis. However, currently over 50% of the *Shigella*-associated diarrhea cases are presenting as watery diarrhea [10, 13–16]. A systematic review and meta-analysis on the identification and management of *Shigella* showed that the sensitivity of dysentery for identifying shigellosis has declined from 85.9% to 1.9% over time [17]. WHO diarrhea treatment guidelines recommend oral rehydration solution along with the provision of zinc for watery diarrhea cases, keeping antibiotics reserved for severe cholera and cases with dysentery (visible blood or caregiver-reported blood in stool) [18, 19]. Therefore, in the absence of a point-of-care diagnostic assay, this large number of *Shigella*-associated watery diarrhea cases would be undiagnosed and untreated, which poses a severe health risk, including mortality in these children.

We previously developed a rapid LAMP-based diagnostic test (RLDT) for detecting *Shigella* (*ipaH* gene) from stool samples to address this gap in diagnosing shigellosis in low-resource settings [20]. RLDT is simple and rapid (< 1 hour), performed directly from stool with minimum sample preparation, and avoids maintaining a cold chain. RLDT can be performed from fresh stools, frozen stools, rectal swabs, and dry stool spots. Moreover, the analytical lowest detection limit of RLDT is similar to qPCR, and diagnostic sensitivity ranging from 93 to 100% when compared to qPCR [21–23]. The RLDT assay for *Shigella* had been evaluated in India [21], Zambia [23], and Burkina Faso [24].

In this study, we aim to evaluate the feasibility of the RLDT assay for detecting *Shigella* in a rural primary health care setting in Bangladesh for case detection and treatment. We also evaluated the acceptance of RLDT at that health care facility.

## Methods

### Study site

For testing the feasibility of RLDT, we selected a rural primary health care facility, Mirzapur Upazila Health Complex (MUHC), which is a 50-bed sub-district healthcare facility run by the health system of Bangladesh.

Fig 1 illustrates the organizational structure of the health system of Bangladesh and the position of MUHC. The responsibilities of health service delivery are undertaken by two ministries- the Ministry of Health and Family Welfare (MOHFW) and the Ministry of Local Government, Rural Development and Cooperatives - along with their subordinate organizations and service points. Upazila Health Complex, like MUHC, is a part of the primary healthcare system in Bangladesh. MUHC is located in the rural Mirzapur sub-district, 70km northeast of the capital city, Dhaka, of Bangladesh. The facility has a small, basic diagnostic laboratory with two lab personnel. Every year, this sub-district healthcare facility treats around 1,63000 patients, including 6000 diarrheal patients of all ages. Lab personnel of MUHC conduct, on average, 90 laboratory tests every day.

We conducted this study under an ongoing prospective case-control study titled “Impact of Non-Dysentery *Shigella* Infection on the Growth and Health of Children over Time-INSIGHT” [25]. The INSIGHT study enrolled children under five, who were seeking care for diarrhea at the Kumudini hospital (a tertiary care hospital) in Mirzapur and were positive for *Shigella* by the RLDT. These children were followed for one year with home visits twice a week.

**Fig 1.**
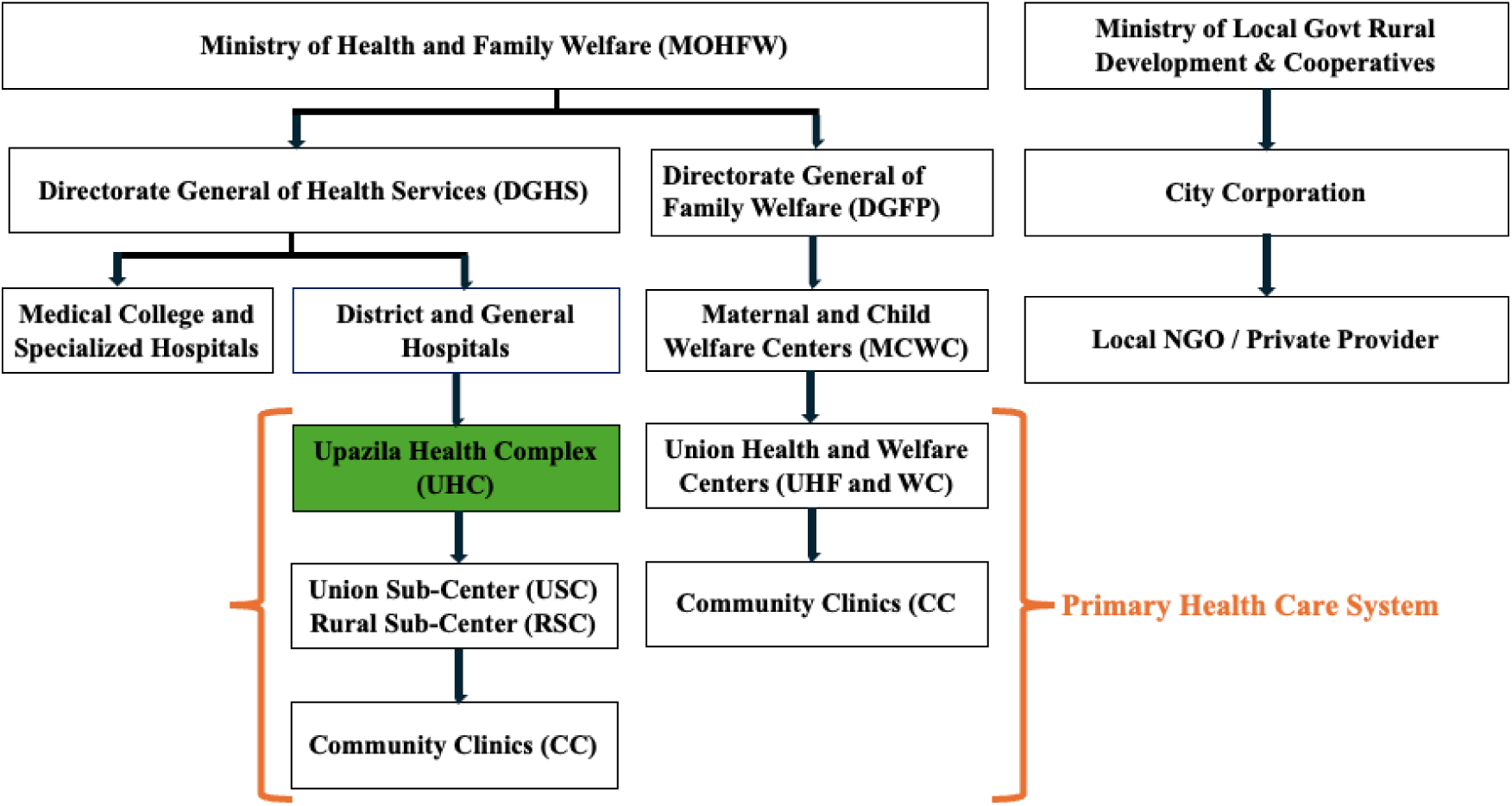
Organogram of the Health System of Bangladesh. The position of the UHCs (eg, MUHC) is shown in green.

### Sample collection and selection for this study

Stool specimens were collected from the acute diarrheal children for screening and enrollment in the INSIGHT study. We also collected stool samples from the enrolled children during the scheduled follow-up visits. All samples were tested by RLDT for detecting *Shigella* in the INSIGHT study laboratory in Mirzapur. If the child with diarrhea was positive for *Shigella*, by RLDT, along with 12% RLDT-*Shigella* negative stool samples were sent to the icddr,b Dhaka clinical laboratory for culture and serotyping. Between 1st February 2023 and 31st October 2023, 228 stool samples were collected and tested with RLDT on the designated days of testing at the MUHC; 137 stool samples were collected from children who were having diarrhea at the time, and the rest were collected during follow-up visits after the diarrhea resolved. These 228 stool samples were used for the RLDT evaluation and feasibility study at the MUHC. The follow-up samples were included to test asymptomatic and *Shigella*-negative samples.

### RLDT test

The details of the RLDT test procedure were described previously [20]. In short, RLDT includes a simple, rapid, and sensitive method for sample preparation directly from stool to facilitate the use of the assay in low-resource settings. Stool samples were added to the sample processing tube with lysis buffer, followed by heat lysis. The processed samples were then added to the lyophilized RLDT reaction strips. Each strip consisted of 8 tubes, organized as two reaction tubes, one for *ipaH*, and one tube for the RLDT assay inhibitor control. The strips were run for 40 minutes in the RLDT reader.

### Study procedure

Two days a week were selected for testing stool samples over a 9-month period in the MUHC. On the testing days, children were enrolled in the INSIGHT study, the stool samples were collected and first tested using RLDT at the INSIGHT laboratory by the INSIGHT study staff, and on the same day, sent to the MUHC for retesting by the MUHC staff. The distance of MUHC is 10 km from the INSIGHT laboratory. At MUCH, the staff registered their results in a logbook at the end of the tests. A total of 54 stool samples were sent for culture to the icddr,b laboratory.

For the evaluation of the acceptability of this assay, we conducted a semi-structured interview among the two lab staff members of MUHC and the Upazila Health and Family Planning Officer (UHFPO), who is the facility head and the lead of the administrative and clinical services of MUHC. Each interview session was conducted in a closed-door setting, so no one person was aware of the responses from another. Each interview session took approximately 25 minutes.

### Ethical consideration

The study protocol was reviewed and approved by the Institutional Review Board of the Johns Hopkins Bloomberg School of Public Health and the Ethical Review Committee of the icddr,b. We obtained informed, voluntary, written consent from the parents/caregivers of the study participants. All the data collection methods were performed in accordance with the Declaration of Helsinki.

### Statistical analysis

The RLDT test results from the MUHC were compared with the INSIGHT study RLDT results (Gold standard) to determine sensitivity and specificity. The test results were run and interpreted by the laboratory personnel of the INSIGHT laboratory and the MUCH, blinded to each other. Sensitivity, specificity, positive predictive value (PPV), negative predictive value (NPV), and test accuracy were expressed as percentages. For measuring the agreement between the tests performed by the study laboratory personnel and the sub-district laboratory personnel, we used Cohen’s Kappa coefficient scoring. The data were analyzed using STATA version 17 (Stata SE V.17, Stata Corp, College Station, Texas, USA).

## RESULTS

### Laboratory set up at the MUHC

The MUHC has a rudimentary laboratory to perform the basic routine diagnostic tests, such as WBC count with ESR, urine RME, Widal test, ASO titres, blood grouping, Rh typing, Serum Glutamate Pyruvate Transaminase (SGPT), Serum Glutamate Oxaloacetate Transaminase SGOT, serum glucose, serum creatinine, serum cholesterol, pregnancy test, etc., of the visiting patients. The laboratory is 330 square feet and equipped with two microscopes, one semi-auto analyzer for biochemical tests, one cell counter, pH meter, two centrifuge machines, rapid checking devices for Dengue, HBsAg, and HIV, reaction checking strips, several micro pipettes, and two refrigerators. RLDT assay requires the RLDT kit (stable at room temperature), a small heat block, and a battery-operated handheld reader for reading the RLDT results. Because of the minimum equipment needed, the RLDT assay was feasible to set up even in this small but crowded laboratory at the MUHC. For RLDT, we set up a small table in a corner of the laboratory for stool receiving and processing activities and set up the RLDT reader on another bench.

### Training of the RLDT assay

Under the INSIGHT study, field laboratories were established. The field laboratory staff were trained by the Johns Hopkins University in RLDT and certified. To conduct the RLDT evaluation and feasibility study, two laboratory personnel of the INSIGHT study were selected as trainers. Both have a Diploma in Medical Technology and have been performing RLDT in the INSIGHT field laboratory for the last 4 years, and have tested over 7000 stool samples with RLDT effectively. The INSIGHT trainers trained the two lab staff at the MUHC who are government employees at the health system of Bangladesh and employed to run the routine laboratory activities at the MUHC. Both of them also have a Diploma in Medical Technology, but neither of them had any previous knowledge or exposure to RLDT or any molecular diagnostic assays, nor had they worked with any stool test. For training, on the first day, a written protocol and a PowerPoint presentation of the procedures were provided to the MUHC staff, followed by hands-on training. MUHC staff then performed the RLDT assay independently but under the observation of the INSIGHT trainers for the next three days, after which they performed the RLDT tests independently without oversight.

### Performance of RLDT in clinical settings

A total of 228 samples, including 137 diarrheal and 91 asymptomatic stool samples, were re-tested by the staff at MUHC. Among these, 41 samples (17.98%) tested RLDT positive by the INSIGHT study staff, and 42 samples (18.42%) tested RLDT positive at the MUHC. The sensitivity, specificity, PPV, NVP, and accuracy of the RLDT test for total samples at MUHC were between 95.2% to 99.5% and for all the asymptomatic samples were between 94.7% to 97.8%(Table 1).

**Table 1.**
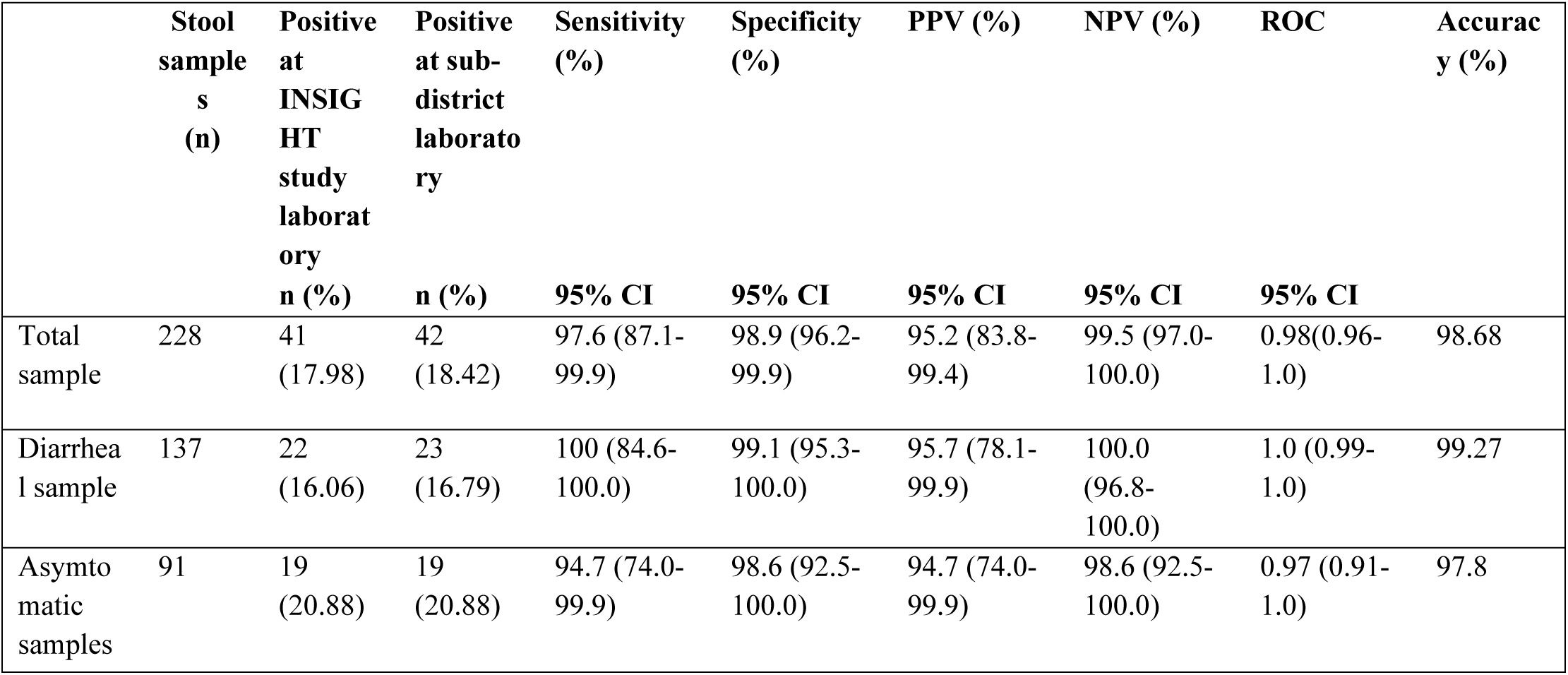
Performance of the tests executed in MUHC laboratory.

|  | Stool<br>sample<br>s<br>(n) | Positive<br>at<br>INSIG<br>HT<br>study<br>laborat<br>ory<br>n (%) | Positive<br>at sub-district<br>laborato<br>ry<br>n (%) | Sensitivity<br>(%)<br>95% CI | Specificity<br>(%)<br>95% CI | PPV (%)<br>95% CI | NPV (%)<br>95% CI | ROC<br>95% CI | Accurac<br>y (%) |
| --- | --- | --- | --- | --- | --- | --- | --- | --- | --- |
| Total<br>sample | 228 | 41<br>(17.98) | 42<br>(18.42) | 97.6 (87.1-99.9) | 98.9 (96.2-99.9) | 95.2 (83.8-99.4) | 99.5 (97.0-100.0) | 0.98(0.96-1.0) | 98.68 |
| Diarrhea<br>l sample | 137 | 22<br>(16.06) | 23<br>(16.79) | 100 (84.6-100.0) | 99.1 (95.3-100.0) | 95.7 (78.1-99.9) | 100.0<br>(96.8-100.0) | 1.0 (0.99-1.0) | 99.27 |
| Asympto<br>matic<br>samples | 91 | 19<br>(20.88) | 19<br>(20.88) | 94.7 (74.0-99.9) | 98.6 (92.5-100.0) | 94.7 (74.0-99.9) | 98.6 (92.5-100.0) | 0.97 (0.91-1.0) | 97.8 |

The agreement between the performance of MUHC staff and the INSIGHT laboratory staff in RLDT testing was highly consistent, achieving a Kappa coefficient score of 0.9558. (Fig 2)

**Fig 2.**
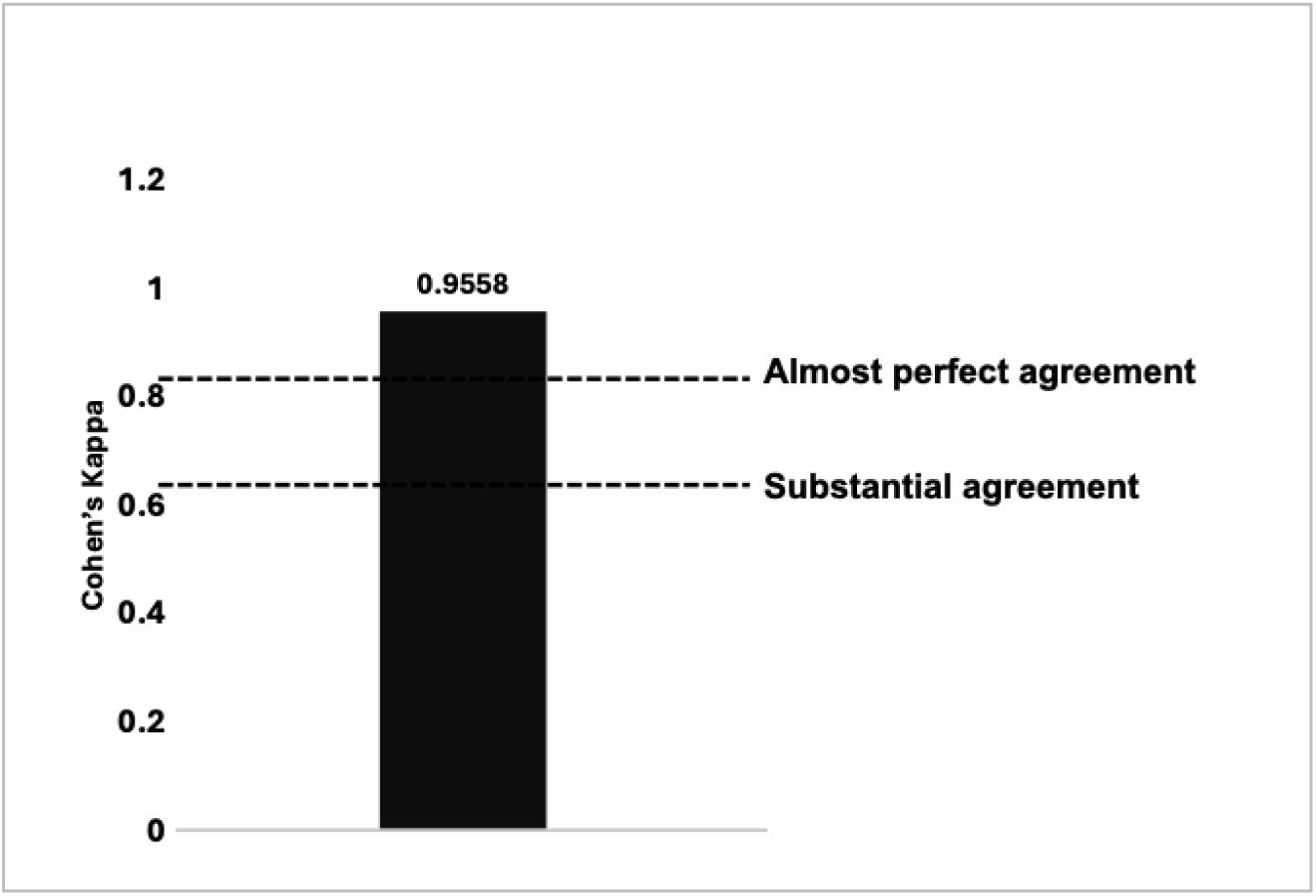
Agreement between the performance of the RLDT conducted by MUHC personnel and INSIGHT personnel.

### Comparison of RLDT with culture

Among the 228 stool samples, all the RLDT-positive (by INSIGHT laboratory) diarrheal stool samples (n=22), 20% of the RLDT-negative diarrheal stool samples (n=23), and nine non-diarrheal RLDT-positive samples were sent for stool culture. From the RLDT positive diarrheal samples, 9 (40.9%), and from the non-diarrheal samples, 2 (22.22%), were culture positive for *Shigella* spp. None of the RLDT negative stool samples yielded any *Shigella* isolates by culture.

### Qualitative Evaluation

The two medical technologists and the Upazila Health and Family Planning Officer (UHFPO), of MUHC were asked for their perception of RLDT and the benefits and challenges of implementing RLDT at the subdistrict level of the health system. Both medical technologists were receptive to RLDT’s potential to support treatment in underserved communities. Both acknowledged that the assay is simple and can be performed at their facility, although they differed on their opinion on the time required to perform the RLDT. While one of them thinks ∼50 minutes for the whole assay with the hands-on time of ∼6 minutes is reasonable and acceptable, the other said that although he could utilize the 40 minutes that was the reaction time in the reader for other work, a shorter duration would be better. According to both, if the higher authority (UHFPO) agrees and supports including RLDT in diagnostic algorithms at the hospitals, then scaling up RLDT across health facilities is possible. Both expressed concerns regarding the challenges of limited human resources, short duty hours, and a considerable workload in attending to the testing needs of a large patient population.

The UHFPO, with four years of experience as head of MUHC, also viewed RLDT positively, recognizing its role in improving diarrhea diagnosis and treatment decision-making and linking it to antimicrobial resistance (AMR) mitigation. To him, RLDT is feasible to deploy in health facilities, with no significant staff-reported challenges noted. He mentioned that initially, the medical technologists were reluctant to absorb this extra workload; however, after starting the RLDT test, they were willing and cooperative. He also added that as government employees, the medical technologists have to follow the directives of the authority. While acknowledging that RLDT is simple and can be scaled up, he also emphasized the need for improved lab conditions, more hands-on training on assays, and the absence of a system for quality monitoring and compliance, which are critical gaps for sustained implementation of any diagnostic test.

### Challenges observed by the study investigators

MUHC is one of the busiest sub-district healthcare facilities with the highest patient turnover in that area. This is the only facility with a government-funded laboratory in that locality, which provides testing at subsidised rates. Thus, the laboratory personnel were often occupied with the routine laboratory tests to meet the need for the enormous patient load. Hence, even the technologists were willing to do the RLDT test, since the RLDT was not included in their scheduled tests, sometimes, absorbing the extra time to perform the RLDT test was challenging.

## DISCUSSION

This is the first study to evaluate the feasibility and applicability of RLDT for the detection of *Shigella* spp. in a health care facility. This study demonstrated that the RLDT could be performed in a primary health care facility in rural Bangladesh. This study also established that the Government lab staff members of a rural basic health facility, who had no prior experience in molecular tests or handling stool samples, could be trained to perform RLDT. In addition, since RLDT is simple and requires minimal equipment, it was feasible to perform in a rudimentary, small clinical laboratory of the MUHC.

The performance of the RLDT assay in detecting *Shigella* at MUHC was comparable to the performance of the RLDT by the experienced INSIGHT lab personnel, with a sensitivity and specificity of over 97%. There was almost perfect agreement between the tests performed by the MUHC personnel and the INSIGHT personnel. Similar high sensitivity and specificity of the MUHC RLDT tests were also found when analyzed within the diarrhea and non-diarrheal stool samples. Of note, all the culture-positive stool samples were also positive by RLDT, and none of the RLDT-negative stool samples were positive by culture, which supports the high specificity of the RLDT assay. The diagnostic performance of RLDT for detecting *Shigella* has been field evaluated and reported similar observations in India, Zambia, and Burkina Faso [21–23]. These studies compared RLDT with culture, PCR, and quantitative PCR (qPCR) methods, which resulted in a sensitivity and specificity of RLDT for detecting *Shigella* compared to qPCR ranging from 93 to 100%.

As shown in the previous studies, the sensitivity and the lowest detection limit of RLDT were comparable with qPCR and more than two-fold higher than culture [21–25]. Our study also showed similar results, that only 41% of the RLDT-positive diarrheal stool samples were culture positive. A reanalysis of GEMS data reported that culture missed half of the *Shigella* attributed MSD diarrhea cases and also missed half of the deaths caused by *Shigella* [10, 16, 17]. The implication of this is that a culture, like a low-sensitivity assay, may miss many shigellosis cases, which, if treated, might be life-saving [10, 16], and the RLDT assay, which is both simple and sensitive, could fill this gap.

The qualitative interviews of the medical technologists and the higher authority at the MUHC highlighted both the potential benefits and key challenges of implementing RLDT at the subdistrict level. While they acknowledged RLDT as a simple assay that could be performed at the MUHC with limited capacity and scaled up, they also highlighted the challenges of low human resources and a considerable workload, which is common in these healthcare facilities. However, they also mentioned that if RLDT testing is included in the policy guidelines for diarrhea diagnosis and treatment, it could be implemented.

Diarrhea can be caused by an array of bacteria, viruses, or parasites. The clinical features of diarrhea alone cannot distinguish between bacterial and viral diarrhea. The epidemiology of *Shigella* has changed over time, and there are more watery diarrhea shigellosis cases than *Shigella* dysentery. Evidence is generating that the treatment of watery diarrhea shigellosis may save lives and reduce long-term disabilities, which include stunting [26]. However, antibiotic treatment will require confirming shigellosis before the treatment decision. Irrational use of antibiotics contributes to the development and spread of antimicrobial resistance, both at the individual level and at the population level. Our recent study conducted on 7000 children in Bangladesh revealed that 55% of the children with watery diarrhea received antibiotics, and 77% of them received antibiotics irrationally from a local drugstore without presenting any prescription from a registered doctor [27]. Therefore, to treat watery diarrhea shigellosis and better diagnose dysentery cases, a simple and sensitive assay for detecting *Shigella* is critical. This study demonstrated that RLDT is feasible for application in healthcare facilities to detect shigellosis and thus has the potential to fill the void. RLDT could also reduce the overuse of antibiotics and thus reduce antimicrobial resistance.

RLDT is also developed and field tested for the detection of enterotoxigenic *E. coli* and *Vibrio cholerae*, and the development of RLDT tests to detect other pathogens is underway. Therefore, once a health facility is trained in RLDT and has the RLDT reader, it can detect all these pathogens. RLDT is a semiquantitative assay, and the time to reach the assay threshold has a linear relation to the CFU of the target pathogen in the stool [20]. Therefore, to identify cases that are attributed to *Shigella*, a threshold could be set up in the RLDT reader to determine positive or negative. A study to determine the threshold is underway.

### Strengths and limitations

This study was conducted in a sub-district health facility of rural Bangladesh, which represents the primary health care system of Bangladesh. Laboratory personnel trained in RLDT belonged to the health system of Bangladesh. This study also has limitations. This was a pilot study and was therefore only conducted in a single healthcare facility, and the qualitative study was focused only on the personnel relevant to this facility. In the future, RLDT should be evaluated at various health facility levels within a country and on different continents, as the challenges may vary by the type of facility and location.

## Conclusion

RLDT could be implemented in a rural primary health care facility. RLDT is a potential diagnostic tool to meet the challenges of the diagnosis of shigellosis for evidence-based treatment to reduce shigellosis morbidity and mortality, as well as reduce antimicrobial resistance. Large-scale adoption would enable prompt identification and management of shigellosis in low-resource settings and would also help in understanding the true burden of this disease.

## Data availability

All data generated or analysed during this study are included in this article. Any additional data related to this paper is available upon request to the corresponding author

## Acknowledgement

We thank the staff members of MUHC for their cooperation and contribution in testing samples for the study. We thank all the staff of the INSIGHT study for their hard work and invaluable contributions. We also thank all the children and their caregivers who participated in this study. We acknowledge the contribution of icddr,b’s core donors, including the Government of Bangladesh, and Canada, for their continuous support and commitment to icddr,b’s research efforts.

## Author contributions

**Conceptualization:** S.C, S.D., and A.S.G.F.;

**Methodology:** S.D., S.C., A.S.G.F., and K.F.;

**Investigation:** S.D., E.S., A.S.G.F., S.C., M.A., M.R.R., M.M.R., S.R.D., and F.N.,

**Writing—original draft preparation:** S.D;

**Writing, review and editing:** S.D., S.C., A.S.G.F., K.F.; F.N.; and T.A.;

**Funding acquisition:** S.C.

All authors have reviewed and agreed to the published version of the manuscript.

## References

1. Anderson, J.D.t., et al., Burden of enterotoxigenic Escherichia coli and shigella non-fatal diarrhoeal infections in 79 low-income and lower middle-income countries: a modelling analysis. Lancet Glob Health, 2019. 7(3): p. e321–e330.

2. Khalil, I.A., et al., Morbidity and mortality due to shigella and enterotoxigenic Escherichia coli diarrhoea: the Global Burden of Disease Study 1990-2016. Lancet Infect Dis, 2018. 18(11): p. 1229–1240.

3. Murray, C.J., et al., *Global, regional, and national disability-adjusted life years (DALYs) for 306 diseases and injuries and healthy life expectancy (HALE) for 188 countries*, *1990-2013: quantifying the epidemiological transition*. Lancet, 2015. 386(10009): p. 2145–91.

4. Murray, C.J., et al., *Disability-adjusted life years (DALYs) for 291 diseases and injuries in 21 regions*, *1990-2010: a systematic analysis for the Global Burden of Disease Study 2010*. Lancet, 2012. 380(9859): p. 2197–223.

5. WHO, Shigella (https://www.who.int/teams/immunization-vaccines-and-biologicals/diseases/shigella). 2022.

6. Rogawski McQuade, E.T., et al., Epidemiology of <em>Shigella</em> species and serotypes in children: a&#xa0;retrospective substudy of the MAL-ED observational birth cohort study. The Lancet Microbe.

7. Bagamian, K.H., et al., Heterogeneity in enterotoxigenic Escherichia coli and shigella infections in children under 5 years of age from 11 African countries: a subnational approach quantifying risk, mortality, morbidity, and stunting. Lancet Glob Health, 2020. 8(1): p. e101–e112.

8. Bagamian, K.H., et al., Shigella and childhood stunting: Evidence, gaps, and future research directions. PLoS Negl Trop Dis, 2023. 17(9): p. e0011475.

9. Lindsay, B., et al., Quantitative PCR for detection of Shigella improves ascertainment of Shigella burden in children with moderate-to-severe diarrhea in low-income countries. J Clin Microbiol, 2013. 51(6): p. 1740–6.

10. Liu, J., et al., Use of quantitative molecular diagnostic methods to identify causes of diarrhoea in children: a reanalysis of the GEMS case-control study. Lancet, 2016. 388(10051): p. 1291–301.

11. Tickell, K.D., et al., Identification and management of Shigella infection in children with diarrhoea: a systematic review and meta-analysis. Lancet Glob Health, 2017. 5(12): p. e1235–e1248.

12. Rogawski, E.T., et al., Use of quantitative molecular diagnostic methods to investigate the effect of enteropathogen infections on linear growth in children in low-resource settings: longitudinal analysis of results from the MAL-ED cohort study. Lancet Glob Health, 2018. 6(12): p. e1319–e1328.

13. Lindsay, B., et al., Association Between Shigella Infection and Diarrhea Varies Based on Location and Age of Children. Am J Trop Med Hyg, 2015. 93(5): p. 918–24.

14. Kotloff, K.L., et al., Burden and aetiology of diarrhoeal disease in infants and young children in developing countries (the Global Enteric Multicenter Study, GEMS): a prospective, case-control study. Lancet, 2013. 382(9888): p. 209–22.

15. Kotloff, K.L., et al., The incidence, aetiology, and adverse clinical consequences of less severe diarrhoeal episodes among infants and children residing in low-income and middle-income countries: a 12-month case-control study as a follow-on to the Global Enteric Multicenter Study (GEMS). Lancet Glob Health, 2019. 7(5): p. e568–e584.

16. Kotloff, K.L., et al., Global burden of Shigella infections: implications for vaccine development and implementation of control strategies. Bull World Health Organ, 1999. 77(8): p. 651–66.

17. Miti, S., et al., Sensitivity and predictive value of dysentery in diagnosing shigellosis among under five children in Zambia. PLoS One, 2023. 18(2): p. e0279012.

18. Organization, W.H., Guidelines for the Control of Shigellosis, Including Epidemics Due to Shigella Dysenteriae Type 1 ( https://www.who.int/publications/i/item/9241592330). 2005.

19. Organization, W.H., Pocket Book of Hospital Care for Children ( https://www.who.int/publications/i/item/978-92-4-154837-3). 2013. 2nd edition.

20. Chakraborty, S., S. Connor, and M. Velagic, Development of a simple, rapid, and sensitive diagnostic assay for enterotoxigenic E. coli and Shigella spp applicable to endemic countries. PLoS Negl Trop Dis, 2022. 16(1): p. e0010180.

21. Chowdhury, G., et al., Field evaluation of a simple and rapid diagnostic test, RLDT to detect Shigella and enterotoxigenic E. coli in Indian children. Sci Rep, 2024. 14(1): p. 8816.

22. Connor, S., et al., Evaluation of a simple, rapid and field-adapted diagnostic assay for enterotoxigenic E. coli and Shigella. PLoS Negl Trop Dis, 2022. 16(2): p. e0010192.

23. Silwamba, S., et al., Field evaluation of a novel, rapid diagnostic assay, and molecular epidemiology of enterotoxigenic E. coli among Zambian children presenting with diarrhea. PLoS Negl Trop Dis, 2022. 16(8): p. e0010207.

24. Héma, A., et al., Contribution of the Rapid LAMP-Based Diagnostic Test (RLDT) to the Evaluation of Enterotoxigenic Escherichia coli (ETEC) and Shigella in Childhood Diarrhea in the Peri-Urban Area of Ouagadougou, Burkina Faso. Microorganisms, 2023. 11(11).

25. Chakraborty, S., et al., The Impact of Non-Dysentery Shigella Infection on the Growth and Health of Children over Time (INSIGHT)-A Prospective Case-Control Study Protocol. Microorganisms, 2024. 12(8).

26. Cornick, J., et al., Azithromycin treatment response as a probe to attribute bacterial etiologies of diarrhea using molecular diagnostics: a reanalysis of the Antibiotics for Children with Severe Diarrhea (ABCD) trial. Front Microbiol, 2025. 16: p. 1606207.

27. Dash, S., et al., Healthcare seeking behavior and antibiotic use for diarrhea among children in rural Bangladesh before seeking care at a healthcare facility. Sci Rep, 2025. 15(1): p. 26359.

